## Supplementary file 1 for "Who does tracing work for? Characteristics of clients successfully re-engaged in ART care in sub-Saharan Africa after a tracing intervention: a systematic review"

NIHR | National Institute for Health and Care Research

PROSPERO
International prospective register of systematic reviews

### Who does tracing work for? Characteristics of clients successfully re-engaged in ART care in sub-Saharan Africa after a tracing intervention: a systematic review

*Allison Morgan, Sydney Rosen, David Flynn, Mariet Benade, Linda Sande, Anushka Marri*

Review methods were amended after registration. Please see the revision notes and previous versions for detail.
To enable PROSPERO to focus on COVID-19 submissions, this registration record has undergone basic automated checks for eligibility and is published exactly as submitted. PROSPERO has never provided peer review, and usual checking by the PROSPERO team does not endorse content. Therefore, automatically published records should be treated as any other PROSPERO registration. Further detail is provided here.

**Citation** 1 change
Allison Morgan, Sydney Rosen, David Flynn, Mariet Benade, Linda Sande, Anushka Marri. Who does tracing work for? Characteristics of clients successfully re-engaged in ART care in sub-Saharan Africa after a tracing intervention: a systematic review. PROSPERO 2024 Available from https://www.crd.york.ac.uk/PROSPERO/view/CRD42024534323

#### REVIEW TITLE AND BASIC DETAILS

##### Review title
Who does tracing work for? Characteristics of clients successfully re-engaged in ART care in sub-Saharan Africa after a tracing intervention: a systematic review

##### Original language title
English

##### Review objectives
*What are the characteristics of ART clients who experience a treatment interruption or disengage from care and are successfully traced and return to care following an active in-person or electronic tracing intervention?*

A standard intervention for improving continuous engagement in HIV treatment programs, tracing aims to contact clients who are late for scheduled clinic visits through either electronic (e.g. telephone) or in-person (e.g. home visit) interactions. Most policies and guidelines for tracing interventions prioritize clients' clinical condition and clinical consequences of disengagement from care, such that those who are at greater risk of negative health outcomes are traced first. It is likely, however, that individual demographic, socioeconomic, behavioral, and other characteristics not associated with clinical condition predict whether or not the tracing intervention will be successful (i.e. the patient will re-engage in care following tracing). This review will consider if any reported characteristics predict tracing success and could thus be incorporated into prioritization algorithms for tracing interventions (i.e., "for whom does tracing work and lead to re-engagement in care?").

**Keywords**
antiretroviral therapy, HIV, Systematic review, tracing, tracking

#### SEARCHING AND SCREENING

##### Searches

We will search PubMed, Embase, and Web of Science. We will manually search reference lists from sources identified in the primary search to identify additional relevant sources. We will also search major international HIV conference (CROI, IAS) abstracts. The search will be limited to studies with a majority of data generated between January 1, 2004 and the earliest date on which one of the electronic indices listed above is searched. Searches will be limited only to those printed in English and to studies conducted in sub-Saharan Africa. The search will be rerun prior to publication to identify newly published work.

##### Study design

We will include studies that report characteristics of traced clients, stratified by whether they re-engaged with care and/or could be reached. We expect these to be mostly interventional studies. Qualitative studies will be excluded unless there is sufficient information about client demographics, stratified by engagement after tracing.

#### ELIGIBILITY CRITERIA

##### Condition or domain being studied

Re-engagement in antiretroviral therapy among clients with HIV who have been lost to follow-up

##### Population

Inclusion: Adults (over 18 years of age) with HIV who have disengaged from care and were traced through an in-person or electronic intervention for re-engagement in care, using primary papers' authors' definitions of disengagement and tracing.
Exclusion: Adolescents and children (under 18 years of age); pregnant and postpartum women who are receiving care through ANC or MCH clinics.

##### Intervention(s) or exposure(s)

Active in-person or electronic follow up by professional, lay, or volunteer healthcare workers with the aim of re-engagement in ART is the intervention of interest.

##### Comparator(s) or control(s)

We will observe differences in client characteristics for those who re-engaged with care as a result of tracing, compared to those who did not re-engage after tracing and with those who could not be reached by the tracing intervention.
Acknowledging that only about half of individuals targeted by tracing interventions can be reached, we will also attempt to describe characteristics (and differences) among clients who are reached, compared to those who cannot be reached.

##### Context

Studies conducted in sub-Saharan Africa (World Bank classification). We will include studies from a variety of settings, including rural and urban contexts, and will record setting descriptions as possible.

#### OUTCOMES TO BE ANALYSED

##### Main outcomes

All reported characteristics of the three main groups (traced and returned, traced but not returned, could not be found for tracing), including sex, age, setting (rural/urban/periurban), distance from clinic, socio-economic status, HIV treatment history, household characteristics, behavioral characteristics, self-reported preferences and concerns, and any other characteristics reported by the original studies.

##### Additional outcomes

None

#### DATA COLLECTION PROCESS

##### Data extraction (selection and coding)

Rayyan will be used to screen and manage study inclusion and exclusion. References will be managed using Mendeley. Two independent reviewers will screen titles and abstracts, conduct full-text screening for outcomes of interest, and extract data. In instances of disagreements in inclusion/exclusion, a third co-author will be asked to review the source in question.
Two independent reviewers will extract the following variables from each article included after full-text screening:
Study design (intervention, observational)
Country
District/municipality
Facility type
Rural/urban setting
Study period/date
Sample size
Tracing intervention (description of activity)
Criteria for triggering tracing (number of days late for scheduled visit, clinical condition, other prioritization considerations, etc.)

Cadre carrying out tracing intervention (nurses, community health workers, etc.)
Intervention specified in national guidelines v study-, facility-, or partner-specific intervention
Tracing trigger for each participant
Number of tracing efforts attempted/participant
Proportion of target clients reached by tracing intervention
Proportion of clients re-engaged after tracing interventions
Time to re-engagement after tracing interventions
Client characteristics, stratified by tracing outcome
Age
Sex
Clinical status (CD4 count, symptoms, etc.)
Socioeconomic indicators (income, employment, housing, etc.)
Household characteristics (including others with HIV/on ART)
Residential location history (mobility/migration)
Distance to clinic
HIV history/treatment
Reasons for disengagement/interruption
Self-reported concerns and preferences (e.g. stigma, experience with providers, etc.)
Any other characteristics reported in source studies

##### Risk of bias (quality) assessment

All sources and variables will be evaluated/extracted by investigators independently to reduce bias. We conduct the review using principals from the Joanna Briggs Institute checklist, by conducting a comprehensive search (including searching conference abstracts) to minimize publication bias; piloting the search strategy to ensure that the independent reviewers have a high degree of agreement; ensuring that the dissemination of findings are reflective of the results of the literature review; and further adhering to the guidelines of the checklist.

#### PLANNED DATA SYNTHESIS

##### Strategy for data synthesis

We will present descriptive characteristics for each article. We will report on the following outcomes as are available: description and characteristics of the tracing interventions; proportion of clients able to be traced; proportion of clients returned to care after tracing; and compare client demographics between clients returned to care and those who remained lost to follow-up. Pooled analysis will be conducted for studies that report on similar interventions and outcomes; however, we anticipate a high degree of heterogeneity that may preclude pooling results.

##### Analysis of subgroups or subsets

We will analyze characteristics by outcome of tracing intervention. If data allow, we may also stratify by tracing intervention type (in-person vs. electronic) or health facility characteristic.

#### REVIEW AFFILIATION, FUNDING AND PEER REVIEW

**Review team members** 1 change


- Allison Morgan, Department of Global Health, Boston University School of Public Health, Boston, Massachusetts, USA
- Sydney Rosen, Department of Global Health, Boston University School of Public Health, Boston, Massachusetts, USA; Health Economics and Epidemiology Research Office, Wits University, Johannesburg, South Africa
- David Flynn, Alumni Medical Library, Boston University Medical Campus, Boston, Massachusetts, USA
- Mariet Benade, Department of Global Health, Boston University School of Public Health, Boston, Massachusetts, USA
- Linda Sande, Health Economics and Epidemiology Research Office, Wits University, Johannesburg, South Africa
- Anushka Marri, Department of Global Health, Boston University School of Public Health, Boston, Massachusetts


**Review affiliation**

Boston University School of Public Health

**Funding source**

This research is supported by the Bill & Melinda Gates Foundation.

**Named contact**

Sydney Rosen. Department of Global HealthBoston University School of Public Health801 Massachusetts AvenueCrosstown Center, 3rd FloorBoston, MA 02118

#### TIMELINE OF THE REVIEW

**Review timeline**

Start date: 15 April 2024. End date: 31 July 2024

**Date of first submission to PROSPERO**

11 April 2024

**Date of registration in PROSPERO**

22 April 2024

#### CURRENT REVIEW STAGE

**Publication of review results**

The intention is to publish the review once completed.The review will be published in English

**Stage of the review at this submission** 1 change

| Review stage | Started | Completed |
|---|---|---|
| Pilot work | ✓ | ✓ |
| Formal searching/study identification | ✓ | ✓ |
| Screening search results against inclusion criteria | | |
| Data extraction or receipt of IP | | |
| Risk of bias/quality assessment | | |
| Data synthesis | | |

##### Review status
The review is currently planned or ongoing.

#### ADDITIONAL INFORMATION

##### Additional information
None

##### PROSPERO version history

- Version 1.2 published on 20 Jun 2024
- Version 1.1 published on 22 Apr 2024
- Version 1.0 published on 22 Apr 2024

##### Review conflict of interest
None known

##### Country
South Africa, United States of America

##### Other registration details
None

##### Medical Subject Headings
Africa South of the Sahara; Algorithms; Ambulatory Care; Demography; HIV Infections; House Calls; Humans; Outcome Assessment, Health Care; Socioeconomic Factors; Telephone; Treatment Interruption

##### Details of any existing review of the same topic by the same authors
Not applicable

##### Revision note 1 change
The preliminary search is now ongoing, and we have added another researcher, Anushka Marri, to the protocol.

##### Disclaimer

The content of this record displays the information provided by the review team. PROSPERO does not peer review registration records or endorse their content.

PROSPERO accepts and posts the information provided in good faith; responsibility for record content rests with the review team. The owner of this record has affirmed that the information provided is truthful and that they understand that deliberate provision of inaccurate information may be construed as scientific misconduct.

PROSPERO does not accept any liability for the content provided in this record or for its use. Readers use the information provided in this record at their own risk.

Any enquiries about the record should be referred to the named review contact
