## Supplementary file 2 for "Who does tracing work for? Characteristics of clients successfully re-engaged in ART care in sub-Saharan Africa after a tracing intervention: a systematic review"

**Supplementary File 1.** Search Terms

<u>PubMed Search Syntax:</u>

("HIV Infections"[Mesh] OR "HIV Infection*" OR "antiretroviral therapy" OR "ART" OR "antiretrovirals") **AND** ("Sub-Saharan Africa" OR "Southern Africa" OR "Africa, Southern"[MeSH Terms] Africa [MeSH] OR "Africa South of the Sahara"[Mesh] OR "Sub-Saharan Africa" OR "Subsaharan Africa" OR "Africa, Sub-Saharan" OR "Africa South of the Sahara" OR Central-africa* OR Eastern-africa* OR East- africa* OR Southern-africa* OR South-africa* OR Western-africa* OR West-africa* OR Angola* OR Benin OR Botswana* OR "Burkina Faso" OR Burundi* OR Cameroon* OR "Cape Verde" OR "Cabo Verde" OR "Central African Republic" OR Chad* OR Comoros OR Congo* OR DRC OR Brazzaville OR "Cote d'Ivoire" OR "Ivory Coast" OR Djibouti* OR "Equatorial Guinea" OR Eritrea* OR Ethiopia* OR Eswatini* OR Gabon OR Gambia* OR Ghana* OR "Guinea Bissau" OR Guinea* OR Kenya* OR Lesotho* OR Liberia* OR Madagascar OR Malawi* OR Mali* OR Mauritania* OR Mauritius OR Mozambique* OR Namibia* OR Niger* OR Nigeria* OR Rwanda* OR "Sao Tome e Principe" OR Senegal* OR Seychelles OR "Sierra Leone" OR Somalia* OR "South Africa" OR "South Sudan" OR Sudan* OR Swaziland* OR Tanzania* OR Togo* OR Uganda* OR Western Sahara OR Zaire OR Zambia* OR Zimbabwe*) **AND** ("tracing" OR "trace" OR "patient tracing" OR "recall" OR "follow up" or "tracking" OR "reengagement" OR "re-engagement" OR "return to care" OR "Program Evaluation"[Mesh] OR "Program Evaluation" OR "Follow-Up Studies"[Mesh] OR "Follow Up Studies" OR "Follow Up Study" OR "Retention in Care"[Mesh] OR "Retention in Care" OR "Care Retention") **NOT** ("contact tracing") **AND** (2004:2024[pdat])

<u>Embase Search Syntax:</u>
(('human immunodeficiency virus'/exp OR 'hiv' OR 'human immuno deficiency virus' OR 'human immunodeficiency virus' OR 'antiretrovirus agent'/exp OR 'anti retroviral agent' OR 'anti retroviral agents' OR 'anti-retroviral agents' OR 'antiretroviral agent' OR 'antiretroviral agents' OR 'antiretrovirus agent' OR 'antiretroviral therapy'/exp OR 'art (drug therapy)' OR 'anti-retroviral therapy' OR 'antiretroviral therapy') **AND** ('africa'/exp OR 'africa' OR 'eastern africa' OR 'southern africa' OR 'western africa' OR 'africa, eastern' OR 'africa, northern' OR 'africa, southern' OR 'africa, western' OR 'east africa') OR 'central africa'/exp OR 'central africa' OR 'africa, central' OR 'angola'/exp OR 'benin'/exp OR 'botswana'/exp OR 'burkina faso'/exp OR 'burundi'/exp OR 'cameroon'/exp OR 'cape verde'/exp OR 'central african republic'/exp OR 'chad'/exp OR 'comoros'/exp OR 'congo'/exp OR 'democratic republic congo'/exp OR 'cote d`ivoire'/exp OR 'djibouti'/exp OR 'equatorial guinea'/exp OR 'eritrea'/exp OR 'ethiopia'/exp OR 'eswatini'/exp OR 'gabon'/exp OR 'gambia'/exp OR 'ghana'/exp OR 'guinea bissau'/exp OR 'guinea'/exp OR 'kenya'/exp OR 'lesotho'/exp OR 'liberia'/exp OR 'madagascar'/exp OR 'malawi'/exp OR 'mali'/exp OR 'mauritius'/exp OR 'mauritania'/exp OR 'mozambique'/exp OR 'namibia'/exp OR 'niger'/exp OR 'nigeria'/exp OR 'rwanda'/exp OR 'sao tome and principe'/exp OR 'senegal'/exp OR 'seychelles'/exp OR 'sierra leone'/exp OR 'somalia'/exp OR 'south africa'/exp OR 'south sudan'/exp OR 'sudan'/exp OR 'tanzania'/exp OR 'togo'/exp OR 'uganda'/exp OR 'western sahara'/exp OR 'zambia'/exp OR 'zimbabwe'/exp) **AND** ('trace'/exp OR tracing OR 'patient tracing' OR 'recall'/exp OR 'follow up' OR 'follow up study' OR 'follow-up studies' OR 'followup' OR 'lost to follow up' OR 'lost to follow-up' OR 're engagement' OR 'return to care' OR 'program evaluation'/exp OR 'follow up'/exp) **NOT** 'contact tracing' **AND** [embase]/lim NOT ([embase]/lim AND [medline]/lim) **AND** 'human immunodeficiency virus infection'/dm **AND** (2004:py OR 2005:py OR 2006:py OR 2007:py OR 2008:py OR 2009:py OR 2010:py OR 2011:py OR 2012:py OR 2013:py OR 2014:py OR 2015:py OR 2016:py OR 2017:py OR 2018:py OR 2019:py OR 2020:py OR 2021:py OR 2022:py OR 2023:py OR 2024:py)

<u>Web of Science Search Syntax</u>

(TS=(hiv) OR TS=(human immunodeficiency virus) OR TS=(ART) OR TS=(antiretroviral therapy) OR TS=('antiretrovirals')) **AND** (CU=(Africa) OR CU=(Eastern Africa) OR CU=(sub-Saharan Africa) OR CU=(Southern

Africa) OR CU=(Western Africa) OR CU=(sub-Saharan Africa) OR CU=(Africa South of the Sahara) OR CU=(Central Africa) OR CU=(East Africa) OR CU=(South Africa) OR CU=(West Africa) OR CU=(Angola) OR CU=(Benin) OR CU=(Botswana) OR CU=(Burkina Faso) OR CU=(Burundi) OR CU=(Cameroon) OR CU=(Cape Verde) OR CU=(Cabo Verde) OR CU=(Central African Republic) OR CU=(Chad) OR CU=(Comoros) OR CU=(Congo) OR CU=(DRC) OR CU=(Brazzaville) OR CU=(Cote d'Ivoire) OR CU=(Ivory Coast) OR CU=(Djibouti) OR CU=(Equatorial Guinea) OR CU=(Eritrea) OR CU=(Ethiopia) OR CU=(Eswatini) OR CU=(Gabon) OR CU=(Gambia) OR CU=(Ghana) OR CU=(Guinea Bissau) OR CU=(Guinea) OR CU=(Kenya) OR CU=(Lesotho) OR CU=(Liberia) OR CU=(Madagascar) OR CU=(Malawi) OR CU=(Mali) OR CU=(Mauritania) OR CU=(Mauritius) OR CU=(Mozambique) OR CU=(Namibia) OR CU=(Niger) OR CU=(Nigeria) OR CU=(Rwanda) OR CU=(Sao Tome e Principe) OR CU=(Senegal) OR CU=(Seychelles) OR CU=(Sierra Leone) OR CU=(Somalia) OR CU=(South Sudan) OR CU=(Sudan) OR CU=(Swaziland) OR CU=(Tanzania) OR CU=(Togo) OR CU=(Uganda) OR CU=(Western Sahara) OR CU=(Zaire) OR CU=(Zambia) OR CU=(Zimbabwe)) **AND** (ALL=(tracing) OR ALL=(trace) OR ALL=(patient tracing) OR ALL=(recall) OR ALL=(follow up) OR ALL=(tracking) OR ALL=(reengagement) OR ALL=(re-engagement) OR ALL=(return to care) OR ALL=(Program evaluation) OR ALL=(follow-up studies) OR ALL=(follow up studies)) **NOT** (All=(contact tracing))
