## Supplementary file 3 for "Who does tracing work for? Characteristics of clients successfully re-engaged in ART care in sub-Saharan Africa after a tracing intervention: a systematic review"

**Supplementary File 3. Summary of study quality assessment using the Joanna Briggs Institute Critical Appraisal**

| Study ID | Author (year of publication) | Total questions assessed* | Total questions with *no indication* of concern | Proportion of questions with *no indication* of concern | Risk classification |
|---|---|---|---|---|---|
| Kenya 1 | Rebeiro (2017) | 8 | 6 | 75% | Low Risk |
| Malawi 2 | Suffrin (2024) | 8 | 7 | 88% | Low Risk |
| Malawi 1 | Tweya (2010) | 11 | 7 | 64% | Moderate Risk |
| Uganda 1 | Nabaggala (2018) | 11 | 9 | 82% | Low Risk |
| Zambia 1 | Beres (2021) (1) | 11 | 7 | 64% | Moderate Risk |
| Zambia 2 | Beres (2021) (2) | 11 | 8 | 73% | Low Risk |
| Zambia 3 | Krebs (2008) | 11 | 7 | 64% | Moderate Risk |
| South Africa 1 | Njuguna (2024) | 13 | 8 | 62% | Moderate Risk |
| Multi 1 | Bershetyn (2017) | 13 | 9 | 69% | Moderate Risk |

**Tools**

*Tools used included the JBI Critical Appraisal Checklist for Cohort Studies; JBI Critical Appraisal Checklist for Analytical Cross Sectional Studies; and JBI Critical Appraisal Tool for Assessment of Risk of Bias for Randomized Controlled Trials.
